# Researching COVID to enhance recovery (RECOVER) autopsy tissue pathology study protocol: Rationale, objectives, and design

**DOI:** 10.1101/2023.04.27.23289234

**Authors:** Andrea B. Troxel, Marie-Abele C. Bind, Thomas J. Flotte, Carlos Cordon-Cardo, Lauren A. Decker, Aloke V. Finn, Robert F. Padera, R. Ross Reichard, James R. Stone, Natalie L. Adolphi, Faye Victoria C. Casimero, John F. Crary, Jamie Elifritz, Arline Faustin, Saikat Kumar B. Ghosh, Amanda Krausert, Maria Martinez-Lage, Jonathan Melamed, Roger A. Mitchell, Barbara A. Sampson, Alan C. Seifert, Aylin Simsir, Cheryle Adams, Stephanie Haasnoot, Stephanie Hafner, Michelle A. Siciliano, Brittany B. Vallejos, Phoebe Del Boccio, Michelle F. Lamendola-Essel, Chloe E. Young, Deepshikha Kewlani, Precious A. Akinbo, Brendan Parent, Alicia Chung, Teresa C. Cato, Praveen C. Mudumbi, Shari Esquenazi-Karonika, Marion J. Wood, James Chan, Jonathan Monteiro, Daniel J. Shinnick, Tanayott Thaweethai, Amber N. Nguyen, Megan L. Fitzgerald, Alice A. Perlowski, Lauren E. Stiles, Moira L. Paskett, Stuart D. Katz, Andrea S. Foulkes RECOVER Initiative

**Author notes:** These authors contributed equally to this work and are co-lead authors (Andrea B. Troxel, Marie-Abele C. Bind). These authors also contributed equally to this work and are co-senior authors (Stuart D. Katz, Andrea S. Foulkes RECOVER Initiative). Authorship has been determined in accord with ICMJE recommendations. Membership of the RECOVER Initiative is provided in the Acknowledgements. **Funding** This research was funded by the National Institutes of Health (NIH) Agreement OTA OT2HL161847-01 as part of the Researching COVID to Enhance Recovery (RECOVER) research Initiative. **Competing interests** Brendan Parent reports receiving a research gift from United Therapeutics. The remaining co-authors have no competing interest to declare. List of Institutions. Funding Source: NIH 1OT2HL161847-01.

## Abstract

**Importance:** SARS-CoV-2 infection can result in ongoing, relapsing, or new symptoms or organ dysfunction after the acute phase of infection, termed Post-Acute Sequelae of SARS-CoV-2 (PASC), or long COVID. The characteristics, prevalence, trajectory and mechanisms of PASC are poorly understood. The objectives of the Researching COVID to Enhance Recovery (RECOVER) Tissue Pathology Study (RECOVER-Pathology) are to: (1) characterize prevalence and types of organ injury/disease and pathology occurring with PASC; (2) characterize the association of pathologic findings with clinical and other characteristics; (3) define the pathophysiology and mechanisms of PASC, and possible mediation via viral persistence; and (4) establish a post-mortem tissue biobank and post-mortem brain imaging biorepository.

**Methods:** RECOVER-Pathology is a cross-sectional study of decedents dying at least 15 days following initial SARS-CoV-2 infection. Eligible decedents must meet WHO criteria for suspected, probable, or confirmed infection and must be aged 18 years or more at the time of death. Enrollment occurs at 7 sites in four U.S. states and Washington, DC. Comprehensive autopsies are conducted according to a standardized protocol within 24 hours of death; tissue samples are sent to the PASC Biorepository for later analyses. Data on clinical history are collected from the medical records and/or next of kin. The primary study outcomes include an array of pathologic features organized by organ system. Casual inference methods will be employed to investigate associations between risk factors and pathologic outcomes.

**Discussion:** RECOVER-Pathology is the largest autopsy study addressing PASC among US adults. Results of this study are intended to elucidate mechanisms of organ injury and disease and enhance our understanding of the pathophysiology of PASC.

**Clinicaltrials.gov number:** NCT05292274

## Introduction

### Background information

The severe acute respiratory syndrome coronavirus 2 (SARS-CoV-2) is a novel coronavirus strain that emerged at the end of 2019. This strain primarily spreads through aerosols in expiratory gasses in infected individuals with and without symptoms, resulting in the highly contagious coronavirus disease 2019 (COVID-19) and a global pandemic. As of December 2022, approximately 653 million people were infected with COVID-19, with at least 6.7 million deaths globally ^1^.

Ongoing, relapsing or new symptoms and organ dysfunction after the acute phase of SARS-CoV-2 infection (≥30 days) are now termed post-acute sequelae of SARS-CoV-2 infection (PASC), often referred to as Long COVID ^2^. The underlying pathophysiology of persistent symptoms after SARS-CoV-2 infection is unknown but may be attributable to mechanisms such as viral persistence, reactivation of other viruses such as Epstein-Barr virus, vascular endothelial activation and thrombosis, autonomic nerve damage, neuroinflammation in the central nervous system, immune dysregulation including activation of auto-immunity, and/or organ damage caused by hyper-inflammatory response during the acute phase of the disease ^3, 4^. Other contributing causes may include complications of critical illness related to prolonged intubation, prolonged bed rest, and malnutrition, and disruptions of health care access. The role of sociodemographic, clinical, and biological factors is likely to be complex and multifaceted.

This study is part of the NIH Researching COVID to Enhance Recovery (RECOVER) Initiative, which seeks to understand, treat, and prevent the post-acute sequelae of SARS-CoV-2 infection (PASC). For more information on RECOVER, visit https://recovercovid.org/. The goals of the RECOVER tissue pathology study (RECOVER-Pathology) are to identify, evaluate, and characterize the clinical manifestations of PASC by comparing decedents with prior SARS-CoV-2 infection who died with and without PASC and the risk factors associated with the severity of these clinical manifestations. This cross-sectional study focuses on the pathologic manifestations of SARS-CoV-2 infection, while explicitly considering sex as a biological variable and the impact of racial and ethnic disparities on autopsy findings. Data acquired from this study will provide accurate and quantifiable measures for PASC symptoms in selected populations to allow for comparisons among groups and, together with the adult and pediatric cohort studies, will provide insights into mechanisms related to pathogenesis of PASC and its underlying biology.

Autopsies are valuable for elucidating the pathophysiologic mechanisms of novel and poorly understood diseases by enabling deep phenotyping of tissues throughout the body ^5^. Autopsies played a critical role in understanding the pathologic changes in patients dying from acute COVID-19. In this setting, the lungs often showed diffuse alveolar damage and pulmonary thrombi ^6, 7^; extra-pulmonary changes were also identified. Microthrombi and ischemic changes were identified in organs throughout the body ^6, 8–11^. Chronic inflammatory infiltrates and occasionally myocarditis were identified in the heart ^6, 8–11^. Hypoxic-ischemic changes, infarcts, and white matter hemorrhagic lesions were identified in the brain; these may be mediated by immune complexes and complement-activating endothelial cells and platelets ^12–14^. *Ex vivo* magnetic resonance imaging performed on brain tissue of deceased children showed abnormalities such as white matter signal hyperintensity, which corresponded to mineralization^15^. In some patients with acute COVID-19, the peripheral nerves showed neuritis, and the skeletal muscle demonstrated changes such as type 2 atrophy, necrotizing myopathy, and myositis ^16^. Similarly, autopsies on patients with PASC helped elucidate the underlying pathologic changes, specifically whether any of the pathologic features observed during acute infection persisted after the initial infection in survivors. In addition, autopsies provided some answers to the clinical challenges of unexpected and sudden clinical decompensation by showing the macro- and microvascular thrombotic manifestations of the disease and endothelial injury. Scientific investigation of autopsy tissues bolstered the clinical suspicions of cytokine interactions with both the innate and adaptive immune system.^17^

### Study rationale

PASC is a profound public health crisis, one that may be with us for many years, with symptoms that can be incapacitating, prevent people from working, and increase the risk of mortality in both adults and children. As of December 2022, there were over 101 million diagnosed cases of COVID-19 in the United States, certainly an underestimate because of undertesting, underreporting, and home testing, particularly in children ^18, 19^. Given the estimated total number of cases, even a low incidence of PASC would affect millions. The incidence, prevalence, phenotypes, risk factors, mechanisms, and etiology of PASC are currently unknown, limiting opportunities for prevention and treatment ^18^. Therefore, autopsy studies are urgently needed. The RECOVER-Pathology study will enhance the knowledge of the effects of SARS-CoV-2 infection and define and characterize the clinical spectrum of PASC in decedents. Ultimately, we expect these data to elucidate potential mechanisms to inform future preventive and treatment studies.

### Study objectives

The overall scientific objectives of this ongoing study are:

1. To enhance knowledge of the effects of SARS-CoV-2 infection;
2. To define and categorize the clinical spectrum of PASC;
3. To elucidate potential mechanisms to inform future preventive and treatment studies.

The following specific aims are proposed to achieve these objectives.

**Aim 1.** Characterize the prevalence and types of organ injury/disease, pathological features, and distinct phenotypes in decedents with acute SARS-CoV-2 infection (dying 15-60 days after initial infection), and in decedents with prior SARS-CoV-2 infection (dying >60 days after initial infection) with and without symptoms of PASC.

**Aim 2.** Characterize the association of pathological findings with clinical measures of disease severity and associated risk factors in decedents with prior SARS-CoV-2 infection (dying > 60 days after initial SARS-CoV-2 infection) with and without PASC.

*2a.* Characterize the association of COVID-19 disease severity and treatment with pathological findings in decedents with prior SARS-CoV-2 infection with and without PASC.

*2b*. Determine whether pre-infection and peri-infection risk and resiliency factors (e.g., demographic, biological factors, and preexisting clinical comorbidities) are associated with pathological findings in decedents with prior SARS-CoV-2 infection with and without PASC.

**Aim 3.** Define the pathophysiology of and mechanisms associated with pathological findings, including the direct and indirect causal effects of clinical risk factors (e.g., comorbidities, infection severity, hospitalization, intubation, steroids or other treatments) on pathological findings (e.g., inflammation, thrombosis, fibrosis, necrosis) via viral persistence (investigated systematically via molecular methods), in decedents with and without PASC.

**Aim 4.** Contribute biospecimens to the RECOVER Biorepository, including post-mortem tissue samples and post-mortem brain imaging, from decedents with prior SARS-CoV-2 infection, with and without PASC. The primary goal of the brain imaging is to find and characterize lesions in the CNS via image-directed sampling of identified lesions. A secondary goal is to correlate the decedent’s MRI findings with those observed in pre-mortem neuroimaging.

## Methods

### Study design overview

RECOVER-Pathology is a cross-sectional study of decedents enrolled in the tissue pathology cohort at varying stages after infection with SARS-CoV-2 (Figure 1). This study is conducted in the United States and decedents are enrolled through interaction with families of the deceased at inpatient, outpatient, and community-based settings. Study data including demographics, pre-infection laboratory tests and imaging with medical history, vaccination history, acute SARS-CoV-2 infection records, overall health and physical function, and PASC symptoms are collected from the electronic health record (EHR) or from family reports of patient medical history using a case report form.

**Fig 1.**
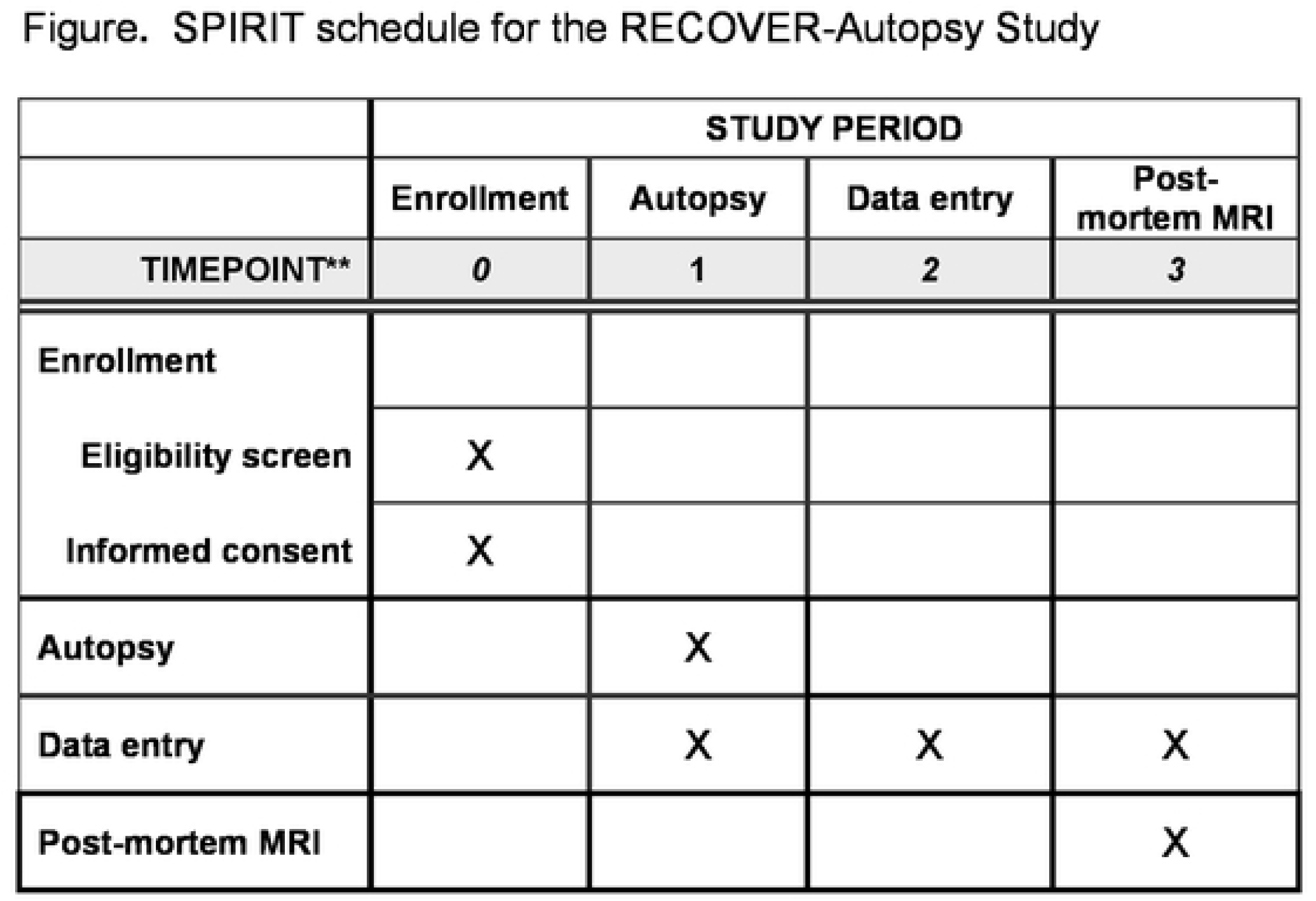
Recruitment Sources for RECOVER-Pathology.

### Setting

RECOVER-Pathology enrollment sites are located in four states (Massachusetts, Minnesota, New Mexico, New York), and the District of Columbia and serve both urban and rural areas with racial and ethnic diversity.

### Study participants

RECOVER-Pathology enrolls decedents after SARS-CoV-2 infection, defined as probable, suspected, or confirmed according to the WHO guideline ^17^, at varying intervals after infection. Some tissue pathology sites are linked with a clinical RECOVER site and may perform autopsies on RECOVER decedents. Additional decedents from extant cohorts and clinical cohorts at these sites and at other RECOVER Initiative tissue pathology cohort sites who are not enrolled in RECOVER, but who have relevant clinical data collected, may be identified for enrollment in the tissue pathology cohort. All RECOVER-Pathology sites select decedents for protocol autopsies according to the inclusion criteria detailed below.

### Cohort subgroups

The RECOVER-Pathology study comprises two distinct subgroups:

#### Post-acute cohort

- Decedents with prior SARS-CoV-2 infection over the age of 18, with and without current or prior PASC-like symptoms, including infected individuals with Multisystem Inflammatory Syndrome in Adults (MIS-A), who die at least 30 days following initial infection.

#### Acute cohort

- SARS-CoV-2 infected decedents who die between 15 and 30 days following initial infection.

If available, relevant retrospective data prior to enrollment are extracted from the electronic health record, existing extant and clinical cohort data, and obtained from next of kin. Decedents contribute to aims according to their SARS-CoV-2 infection/PASC history and progression.

### Inclusion criteria

Decedents aged 18 years and older at the time of death are eligible for inclusion. Previously infected individuals must have suspected, probable, or confirmed SARS-CoV-2 infection as defined by WHO criteria within 24 months of enrollment ^20^. Decedents are eligible regardless of biological sex, race/ethnicity, geography, nationality, severity of disease, underlying health conditions, history of MIS-A or MIS-C (Multisystem Inflammatory Syndrome in Children), or history of SARS-CoV-2 vaccination; those with recurrent and/or post-vaccination (breakthrough) SARS-CoV-2 infections are eligible.

### Classification of PASC

Decedents who die more than 30 days following initial infection are initially categorized as having or not having symptoms of PASC according to the WHO definition ^20^, published by the World Health Organization (WHO) on October 6, 2021. They defined PASC as a condition occurring “in individuals with a history of probable or confirmed SARS-CoV-2 infection, usually 3 months from the onset of COVID-19 with symptoms that last for at least 2 months and cannot be explained by an alternative diagnosis. Common symptoms include fatigue, shortness of breath, cognitive dysfunction, and others and generally have an impact on everyday functioning. Symptoms may be new onset following initial recovery from an acute COVID-19 episode or may persist from the initial illness. Symptoms may also fluctuate or relapse over time.

### Exclusion criteria

Decedents under the age of 18 at the time of death are not eligible. Decedents dying within 14 days of their initial SARS-CoV-2 infection are not eligible. Decedents in whom the postmortem interval (PMI) is greater than 24 hours are not eligible. Decedents with any indications of criminal investigation at time of death are not eligible, nor are those decedents with signs of decomposition and trauma.

### Enrollment targets

To enhance generalizability of findings, a diverse population of decedents will be enrolled. Enrollment is stratified by age group, sex, and race/ethnicity to promote sufficient inclusion of non-Hispanic Blacks, Hispanic/Latinx, Asian Americans, Native Hawaiians, and Pacific Islanders with approximately equal distribution between male and female populations.

### Stratified recruitment

Recruitment will occur across sites (described in section 3.2). Recruitment of SARS-CoV-2 infected decedents will be targeted at:

- 50% female decedents,
- 53% non-Hispanic White, 16% non-Hispanic Black, 27% Hispanic/Latinx, and 4% Asian Americans, Native Hawaiians, Pacific Islanders, American Indians, and Native Alaskan decedents,
- 57% post-acute decedents with PASC, 29% post-acute decedents without PASC, and 14% acute decedents.

Note that the autopsy sites will frequently communicate with RECOVER Clinical Sciences Core (CSC) and Data Resource Core (DRC) to maintain adherence with recruitment targets, and to promote balance in decedents with and without PASC with respect to characteristics (e.g., age, sex, race / ethnicity).

## Study procedures

### Study organizational structure and study management

Scientific leadership for the study and oversight of participating sites is provided by the RECOVER Clinical Science Core (CSC) at the NYU Grossman School of Medicine. The RECOVER CSC collaborates with the RECOVER Data Resource Core at Massachusetts General Hospital for data management and data storage, and the RECOVER Biorepository at Mayo Clinic for biospecimen storage. The activities of the RECOVER cohort studies are overseen by a Steering Committee, an Executive Committee, a RECOVER Senior Oversight Committee, and an Observational Study Monitoring Board (OSMB) that monitors the study data and makes recommendations to correct any problems with the study that may arise. The membership composition for each of these committees can be viewed at https://recovercovid.org/leadership.

### Site selection

RECOVER-Pathology sites were selected by the National Institutes of Health following a competitive application process in May 2021; these sites were invited to participate in Phase I activities in summer 2021. The group worked together to collaboratively develop the draft study protocols, outline the modified gross autopsy procedure, identify primary endpoints, and specify tissues to be sampled. Phase I sites were approved as Phase II enrolling sites by winter 2021.

### Ethical standards and oversight

All procedures conducted as part of this protocol follow the principles outlined in the report of The Committee for the Oversight of Research Involving the Dead.^18^ These include, but are not limited to, dignity, requirement for reasonable scientific validity, confidentiality, consent, non-interference with organ procurement for donation, and protection from harm.

The investigators will ensure that this study is conducted in full conformity with guidance taken from the Code of Federal Regulations (45 CFR 46.102(f)), which states that “research involving deceased persons is not considered human subjects research and does not require IRB oversight, unless the research involves both living and deceased individuals” ^21^.

### Enrollment

To facilitate the difficult conversation with decedents’ legal next of kin concerning participation in RECOVER-Pathology, the CSC developed an information sheet and consent form that describes the study in plain language. Study personnel meet with families to discuss the research and answer any questions, stressing that participation in the study does not prohibit normal funeral and burial practices. Families interested in learning whether a loved one may qualify for the study can evaluate enrollment eligibility at https://recovercovid.org.

The RECOVER-Pathology study also recognizes the importance of engagement with communities and families who may be under-represented in research by partnering with other federal programs such as the Community Engagement Alliance (CEAL), and the Community Engagement Alliance Consultative Resource (CEACR) to extend outreach to a wide variety of potential participants.

### Study follow-up

Upon request, the decedent’s next of kin can receive the standard clinical autopsy report with a final anatomic diagnosis (FAD). This report will normally be available to next of kin within 60 working days. Next of kin do not receive any research results from the RECOVER-Pathology study. However, aggregated study results will be made available.

### Data sources and measurement

Data may be obtained from multiple sources: directly from the decedent’s next of kin, from existing research records (if participating in an ongoing research study, e.g., RECOVER, extant cohorts, clinical cohorts), from the decedent’s electronic health record or existing registries, or from insurance claims data.

Pre-existing EHR data will be extracted electronically where possible or abstracted and recorded on structured electronic case report forms (eCRFs). Prior to data use, a Health Insurance Portability and Accountability Act of 1996 (HIPAA) authorization is obtained from next of kin, subject to the special provisions that apply post-mortem, which allow for special disclosure provisions that are relevant to deceased individuals ^21^. De-identified data collected from any source are recorded in the RECOVER-Pathology study database, with electronic Case Report Forms (eCRFs).

### Study size

The RECOVER-Pathology study aims to enroll 700 decedents, including 100 with acute SARS-CoV-2 infection dying 15-60 days after infection, and 600 with prior SARS-CoV-2 infection (dying >60 days after infection); of these 600, we anticipate 400 with PASC and 200 without PASC. These numbers provide adequate power to evaluate the multiple hypotheses associated with our aims (**Table 2**).

**Table 1.** RECOVER-Pathology Data.

| Operational Data Types | Structured or Unstructured | Examples |
| --- | --- | --- |
| Electronic Health Record (EHR) Data | Unstructured |  |
| EHR data | Structured, complex |  |
| Claims data | Structured, complex |  |
| Biorepository inventory data | Structured, complex | Biospecimens<br>Slides |
| Biorepository asset data | Structured, complex |  |
| Advanced imaging data | Unstructured | Computer Tomography (CT),<br>Magnetic resonance Imaging<br>(MRI) |
| Vaccination status data | Structured |  |
| Physical examination and gross<br>findings | Structured, complex |  |
| Whole slide imaging data | Unstructured | Section cut, stained with H&E<br>and scanned (40X resolution) |
| Autopsy visceral sites digital<br>droplet polymerase chain reaction<br>(ddPCR) for SARS-CoV-2 virus | Structured, complex |  |
| Immunoassays and 'omics data | Structured, complex | Genomics, Proteomics,<br>Transcriptomics,<br>Metabolomics, Meta-<br>genomics microbiome,<br>Epigenetics |

**Table 2.** Sample size for the RECOVER Tissue Pathology Study by PASC status.

|  | PASC status |  | Total |
| --- | --- | --- | --- |
|  | Without PASC | With PASC |  |
| Post-acute decedents (dying > 60 days after SARS-CoV-2 infection) | 200 | 400 | 600 |
| Acute decedents (dying 15-60 days from SARS-CoV-2 infection) | 100 |  | 100 |
| Total | 300 | 400 | 700 |

## Statistical methods

### Power considerations

To address Aim 1, row 6 in Table 3 provides detectable effect sizes and detectable differences in proportions between PASC+ and PASC-decedents for quantitative and dichotomous pathological features. Results are given without and with a multiple comparison adjustment (assuming 50 tests, alpha=0.001 conservative control).

**Table 3.** Detectable effect measures.

| Total sample size (not including acute decedents) | Number of post-acute decedents with PASC | Number of post-acute decedents without PASC | **Detectable standardized effect size** for 80% power, alpha=0.05, 0.001 (i.e., without and with multiple comparison adjustment), based on two independent group, two-sided comparison of means | **Detectable difference in proportion** between groups of post-acute decedents with and without PASC with 80% power, alpha=0.05, 0.001 (i.e., without and with multiple comparison adjustment), based on two independent groups, two-sided comparison of proportions |  |
| --- | --- | --- | --- | --- | --- |
|  |  |  |  | Proportion with feature = 0.2 in PASC-individuals | Proportion with feature = 0.4 in PASC-individuals |
| 300 | 150 | 150 | 0.32, 0.48 | 0.14, 0.22 | 0.16, 0.24 |
| 300 | 200 | 100 | 0.34, 0.51 | 0.15, 0.24 | 0.17, 0.25 |
| 400 | 200 | 200 | 0.28, 0.42 | 0.13, 0.19 | 0.14, 0.21 |
| 400 | 267 | 133 | 0.30, 0.44 | 0.13, 0.20 | 0.15, 0.22 |
| 600 | 300 | 300 | 0.23, 0.34 | 0.10, 0.15 | 0.12, 0.17 |
| 600 | 400 | 200 | 0.24, 0.36 | 0.11, 0.16 | 0.12, 0.18 |

To address Aim 2, a sample size of 400 within one group (e.g., post-acute decedents with PASC) will have greater than 80% power to detect a small effect size, i.e., Cohen’s d=0.28 between individuals with and without the risk factor, assuming 50% of individuals have the risk factor (i.e., 200 with and 200 without the risk factor; row 3 in Table 3).

To address Aim 3 and characterize the detectable effect sizes from the subset of decedents who undergo brain imaging, for example, please see the first row of Table 3. A sample size of 300 provides 80% power to detect a moderate effect size of d=0.32 between individuals with and without the risk factor (within the specified group), assuming 50% of individuals have the risk factor. These calculations are based on a two-group, two-sided comparison of means, and controlling type 1 error at 0.05.

### Methods of data collection

The majority of the study data are collected at autopsy according to the standardized autopsy protocol described in the appendix** These data include structured fields for the gross autopsy findings. The microscopic findings are captured as text fields, and will later be coded into approved terms using SNOMED (Systematized Nomenclature of Medicine) codes; these clinical terminology codes will be finalized following review by the responsible pathologist. Similarly, the FAD will be coded using a standard set of approved terms. Additional EHR and other research data are extracted and entered into eCRFs.

Biospecimen collection is handled in two ways, depending on stability of the sample:

1. Samples for analytes that require rapid freezing are processed locally and sent to the central RECOVER biorepository on dry ice;
2. Other samples are fixed in formalin and shipped directly to the centralized RECOVER biorepository for processing.

### Primary endpoints

Our statistical analyses will examine a number of primary pathological outcomes including, but not limited to, those described in Table 4.

**Table 4:** Primary pathological outcomes.

|  |  |  |
| --- | --- | --- |
| **Cardiovascular system**<br>• fibrosis<br>• non-ischemic inflammation<br>• acute ischemic injury<br>• infarct<br>• atherosclerosis / arteriosclerosis<br>• thrombosis<br>• presence of SARS-CoV-2 | **Lung**<br>• fibrosis<br>• inflammation<br>• thrombosis<br>• presence of SARS-CoV-2 | **Liver**<br>• fibrosis<br>• inflammation<br>• thrombosis<br>• presence of SARS-CoV-2 |
| **Brain**<br>• inflammation<br>• infarct<br>• thrombosis<br>• hemorrhages (non-traumatic)<br>• atherosclerosis / arteriosclerosis<br>• vasculitis<br>• vascular malformation / aneurysm<br>• neurodegenerative disease<br>• presence of SARS-CoV-2 | **Lymph node / spleen**<br>• Hemophagocytosis<br>• reactive lymph nodes<br>• presence of SARS-CoV-2 | **Kidney**<br>• fibrosis<br>• inflammation<br>• thrombosis<br>• presence of SARS-CoV-2 |

### Causal inference methods

We will assess the overlap and balance in covariate distributions (e.g., sex, age, race/ethnicity, hospitalization status) between comparative groups (i.e., acute decedents vs. post-acute decedents without PASC, and post-acute decedents with vs. without PASC) using the empirical distributions of the estimated propensity scores ^22–24^. If there is evidence of covariate imbalance, we will use matched-sampling techniques to improve balance ^22, 25^.

We will test sharp null hypotheses and report unadjusted and adjusted for multiple comparison ^26^ Fisher p-values based on the assumed, hypothetical exposure assignment mechanism implied by the data and the matched-sampling strategy ^24, 27^ Following the advice of Wasserstein et al. ^28^, we will “not base our conclusions solely on whether an association or effect was found to be ‘statistically significant’ (i.e., the p-value passed some arbitrary threshold such as p < 0.05)”, but rather also explore Fisherian intervals ^23^ and consider model-based inference (e.g., Neymanian / Frequentist, Bayesian), i.e., we will define an estimand of interest and provide a point estimate and a corresponding two-sided uncertainty interval (e.g., 95% confidence interval – CI) for that estimand.

### Autopsy protocol

Development of the autopsy protocol was based upon the standard (“hospital”) autopsy practice in which all the major organs are grossly and microscopically evaluated, additional tissue sampling and specimen collection for ancillary testing is performed routinely, and special research protocols are common. Autopsy pathologists with expertise in COVID and PASC worked, along with patient and caregiver representatives, to develop a broad sampling protocol that could address current and future questions about PASC. To ensure specimen integrity across autopsy sites, the postmortem interval (PMI) between death and autopsy is established at 24 hours or less, which includes transportation of the decedent and other elements (e.g., family time with decedent, consenting for autopsy).

A systematic documentation process for the accurate tracking and transfer of specimens to the biobank is critical and was developed collaboratively by the consortium. Upon completion of the autopsy, the samples are logged into a database for tracking and shipping to the PASC Biorepository Core (PBC). Once the autopsy report is finalized, the relevant clinical information and diagnostic findings are entered into a structured database. The database was developed with input and subject matter expertise by the cohort pathologists, working in tandem with the CSC and DRC.

### Biorepository

The centralized PBC was established at the Mayo Clinic to standardize the collection of samples from RECOVER-Pathology, as well as to catalog, store and distribute samples. The PBC provides training in sample collection, a manual of procedures, and kits for supplies with sample return mailing containers. The biospecimen collection process is summarized in Table 5.

**Table 5:** Biospecimen collections.

| **Collection Container** | **# of Container(s)** | **Collection tube Volume (mL)** | **Fraction** | **Aliquot Volume** |
| --- | --- | --- | --- | --- |
| Blood Spot Card | 1 | N/A | Whole Blood | One card with 4 blood spots |
| Serum Separator Tube (SST) | 2 | 8.5 | Serum | 10 x 1 mL |
| Cerebrospinal Fluid (CSF) | 10 | N/A | CSF | 10 x 1 mL |
| Bronchial Swab | 1 | N/A | Cells | Entire tube |
| Stool | 1 | 25 | Stool | 25 mL |
| 56 Tissues<br>(In duplicate) | 112 | Cryosette™ | Snap Frozen Tissue | 17 mm diameter, not to exceed 4 mm in thickness x 2 |
| 56 Tissues | 56 | Standard Tissue Cassette | FFPE Tissue | 2 x 2 x 0.4 cm |

The PBC tissue specimen list for each tissue site sampled in the RECOVER-Pathology protocol is outlined in Table 6.

**Table 6:**
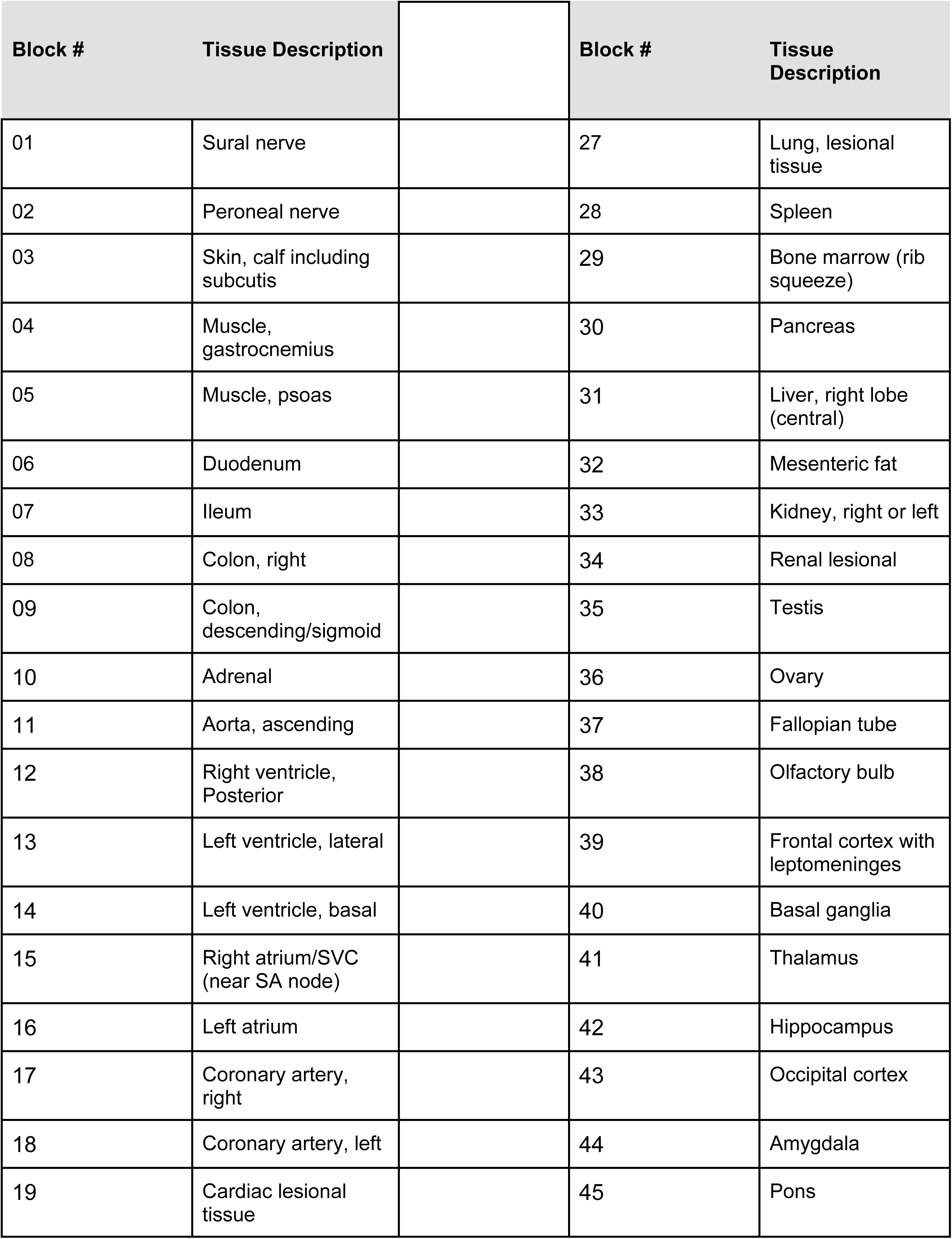
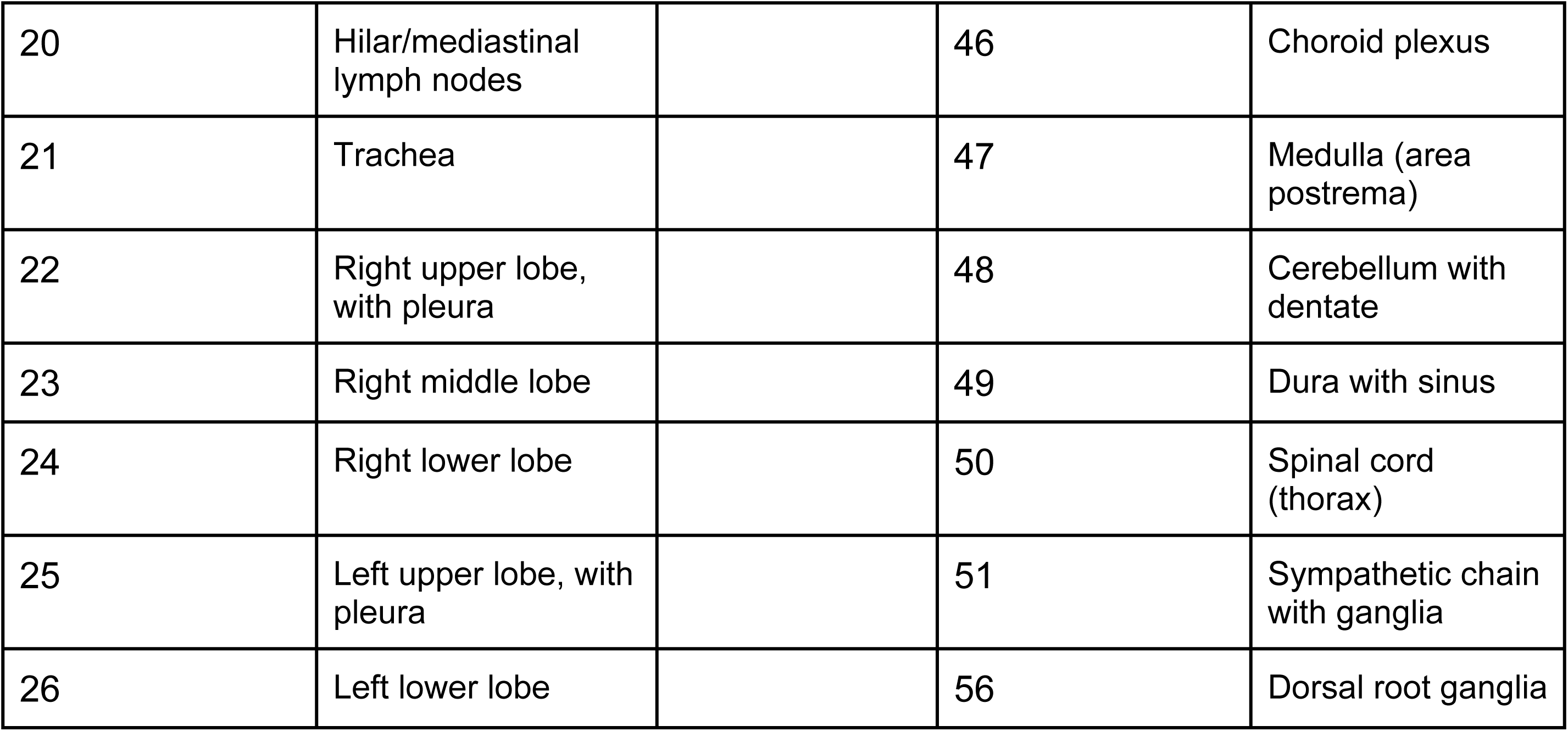
PBC Tissue Specimen Table.

Tissue samples are collected in triplicate, one sample for formalin-fixed, paraffin embedding (FFPE) and two snap-frozen samples. Tissue is collected from fifty-two specified sites (see Figure 2 and Table 6). The autopsy prosectors have the option of selecting four additional tissue sites to document findings at autopsy that are not included in the specified sample sites.

**Fig 2:**
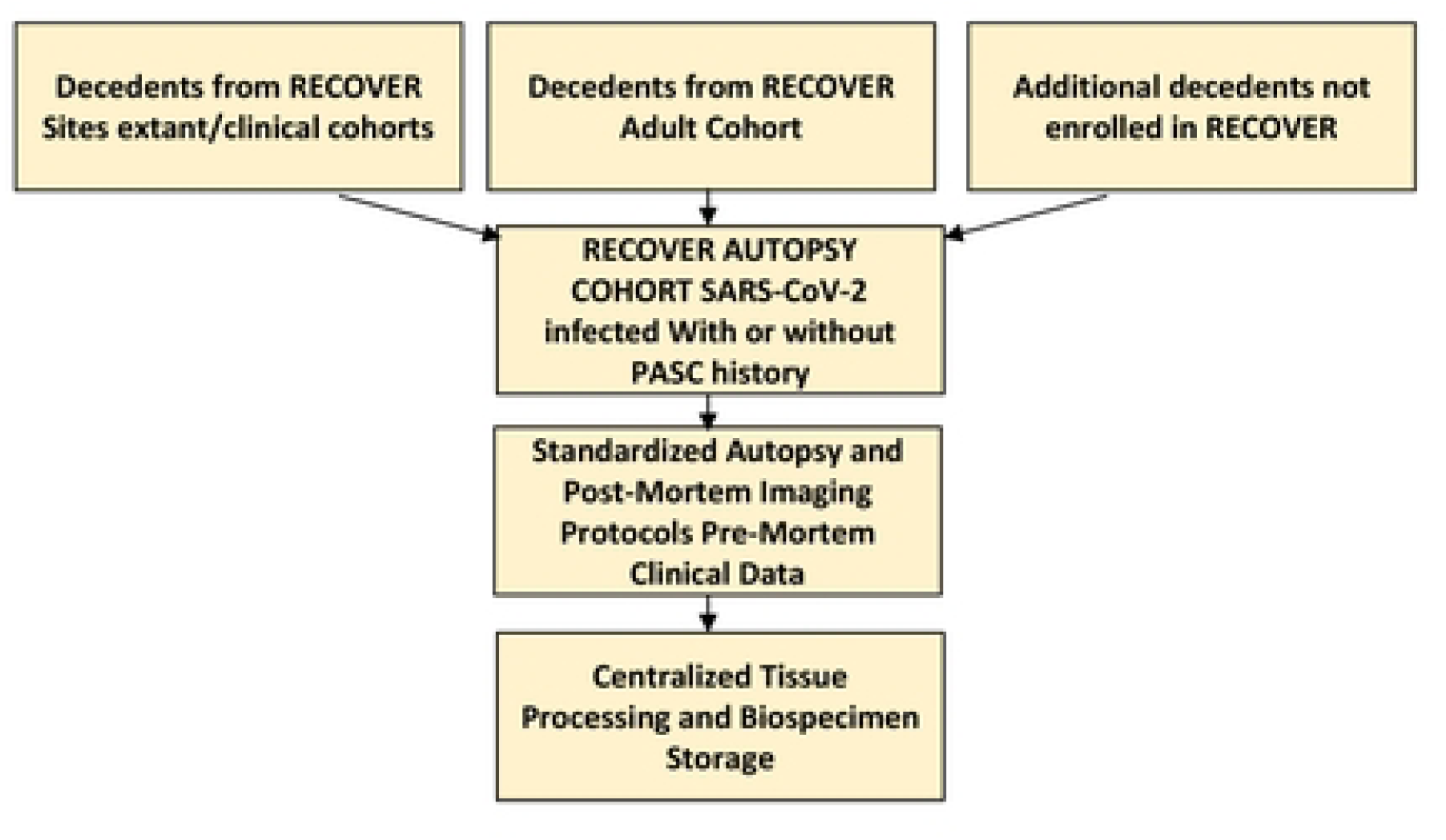
Specified Tissue Collection Sites.

**Fig 3:**
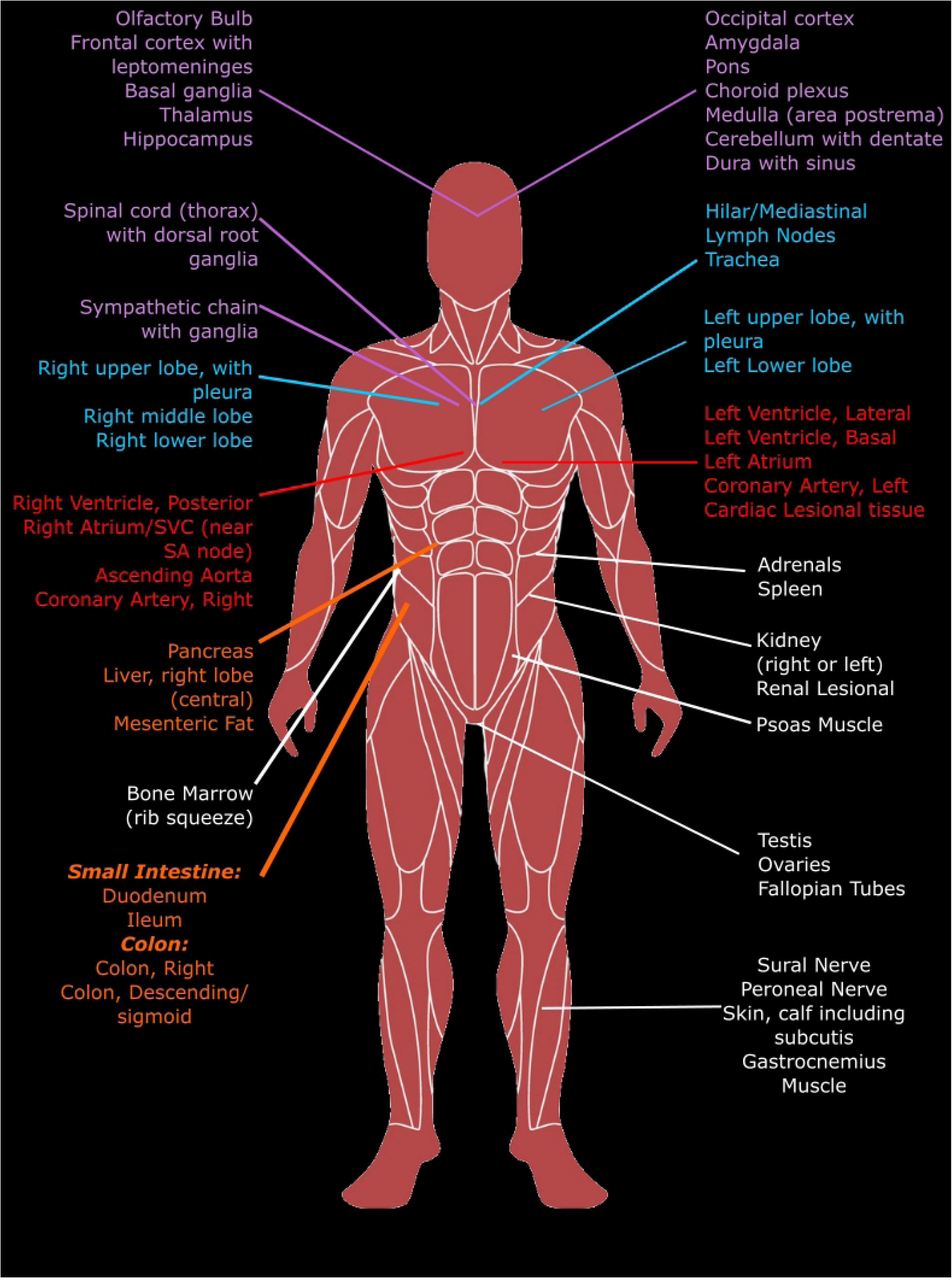

The collection sites process the tissue into FFPE and snap-frozen blocks at their site and send these blocks to the PBC. At the PBC, the blocks are sectioned, stained for hematoxylin and eosin, and the whole slide is imaged at 40x. The slides are reviewed to confirm that they are from the specified sites. The images are transferred to an image repository. The slides and blocks are stored at the PBC.

### Role of the Study Coordinator

In the RECOVER workstream, site coordinators play an important role in screening, consenting, data entry, and assisting the pathology team by conducting specimen procurement procedures. Hundreds of decedents may be screened daily to find qualified candidates. Workflows and alerts developed by the coordinators streamline and facilitate the screening process. Candidates are identified and eligibility is assessed through review of the decedent’s medical history in Epic, other electronic health records (EHRs), or via other available medical records, and coordinators routinely query EHRs for clinical notes and reach out to next of kin to collect as much clinical data as possible. When decedents are deemed eligible for the study, the site coordinator assists the attending clinician with contacting the legal next of kin to obtain consent. The CSC works with site coordinators to facilitate timely and accurate data collection. To ensure that ethical and legal requirements are met, staff are trained on communication and recruitment language strategies. Recruitment materials designed with the goal of informing and comforting the families are provided during the consenting process. Once consent is given, site coordinators arrange transfer of the decedent to the autopsy suite. Each coordinator strives to ensure that the transport process allows the family time with their deceased loved one, while maintaining control of the timeline to ensure compliance with the 24-hour PMI requirement. If any urgent issues occur, staff are prepared to offer support to help address the issue and facilitate enrollment of decedents. The CSC also assists by routine monitoring of enrolling sites, to ensure that sites maintain target goals, good documentation, and operational procedures.

## Discussion

In contrast to most autopsies, the RECOVER-Pathology protocol comprehensively samples tissues with and without any gross or microscopically identifiable pathology. The extensive specimen collection and snap freezing require a team approach, and the efficiency is driven by each team member’s knowledge of the protocol and associated specimen processing. The protocol has been implemented across a range of autopsy practices and will provide a rich resource for future research.

Another distinct feature of this protocol is that the autopsy pathologists routinely seek information regarding SARS-CoV-2 infection from multiple outside sources on all cases. Both medical examiners and hospital-based autopsy pathologists will then seek permission for the study and the autopsy from the decedent’s next of kin. This exemplifies a new and expanding role for autopsy pathologists as monitors of infectious disease throughout the entire population of the recently deceased.

Pathological changes, including persistence of virus within tissues, have been identified in autopsies of patients dying from acute SARS-CoV-2 infection, as well as in patients who have recovered from COVID-19. Persistent viral shedding of SARS-CoV-2 was detected by PCR in the stool up to 210 days after the onset of disease ^30^, in small bowel biopsies up to 173 days from the onset of disease ^31^, in gastrointestinal biopsy specimens 7 months after the onset of disease and associated with gastrointestinal symptoms and symptoms of PASC ^32^. Virus has also been detected by PCR in the cerebrospinal fluid of a patient 114 days after the onset of disease ^33^, and in lung tissue up to 108 days after the onset of disease ^34^. SARS-CoV-2 was found during autopsy in heart tissue by in-situ hybridization and RNA profiling up to 48 days from the onset of disease ^35^. These studies raise the question of whether viral persistence may be an underlying cause of PASC. Previous single-cell RNA profiling from infected lung tissue from autopsies demonstrated that the virus is able to enter diverse cell types ^26^. The RECOVER-Pathology protocol will enable the determination of whether a reservoir of proliferating virus exists in patients with PASC and which tissues and cell types are involved. While a definitive timeline for development of PASC remains elusive, the definition of PASC will be refined in real time by data obtained from all of the RECOVER cohort studies; the RECOVER-Pathology protocol will be amended to reflect these changes. RECOVER-Pathology will make important contributions to our understanding of the mechanisms of PASC, and may lead to additional discoveries and therapeutic interventions.

## Disclaimer statement

This content is solely the responsibility of the authors and does not necessarily represent the official views of the RECOVER Program, the NIH or other funders. This research was funded by the National Institutes of Health (NIH) Agreement OTA OT2HL161847 (contract number 646-501-3654) as part of the Researching COVID to Enhance Recovery (RECOVER) research program.

## Data Availability

No datasets were generated or analysed during the current study. All relevant data from this study will be made available upon study completion.

## Acknowledgements

We would like to thank the National Community Engagement Group (NCEG), all patient, caregiver and community representatives, and the families who gave permission for their loved ones to participate in this autopsy study; we honor the contributions of all participants enrolled in the RECOVER Initiative.

## Supporting information

The supporting information is provided in a supplemental appendix file.

## References

1. Worldometer. (2022). Coronavirus Cases. Retrieved 03 Aug 2022, from https://www.worldometers.info/coronavirus/

2. Callard F, Perego E. How and why patients made Long Covid. Soc Sci Med. 2021 Jan;268:113426. doi: 10.1016/j.socscimed.2020.113426. Epub 2020 Oct 7. PMID: 33199035; PMCID: PMC7539940.

3. Baker SR, Halliday G, Ząbczyk M, Alkarithi G, Macrae FL, Undas A, Hunt BJ, Ariëns RAS. Plasma from patients with pulmonary embolism show aggregates that reduce after anticoagulation. Commun Med (Lond). 2023 Jan 28;3(1):12. doi: 10.1038/s43856-023-00242-8. PMID: 36709220; PMCID: PMC9883810.

4. Nalbandian A, Sehgal K, Gupta A, et al. Post-acute COVID-19 syndrome. Nature Medicine 2021; 27(4): 601–15.

5. Basso C, Stone JR. Autopsy in the Era of Advanced Cardiovascular Imaging. European Heart Journal 2022; 43(26): 2461–8.

6. Bryce C, Grimes Z, Pujadas E, et al. Pathophysiology of SARS-CoV-2: the Mount Sinai COVID-19 autopsy experience. Modern Pathology 2021; 34(8): 1456–67.

7. Jackson NR, Zeigler K, Torrez M, et al. New Mexico’s COVID-19 experience. American Journal of Forensic Medicine and Pathology 2021; 42(1): 1–8.

8. Basso C, Leone O, Rizzo S, et al. Pathological features of COVID-19-associated myocardial injury: A multicentre cardiovascular pathology study. European Heart Journal 2020; 41(39): 3827–35.

9. Rapkiewicz AV, Mai X, Carsons SE, et al. Megakaryocytes and platelet-fibrin thrombi characterize multi-organ thrombosis at autopsy in COVID-19: A case series. EClinicalMedicine 2020; 24:100434.

10. Pellegrini D, Kawakami R, Guagliumi G, et al. Microthrombi as a major cause of cardiac injury in COVID-19: A pathologic study. Circulation 2021; 143(10):1031–42.

11. Bearse M, Hung YP, Krauson AJ, et al. Factors associated with myocardial SARS-CoV-2 infection, myocarditis, and cardiac inflammation in patients with COVID-19. Modern Pathology 2021; 34(7): 1345–57.

12. Lee MH, Perl DP, Steiner J, Pasternack N, Li W, Maric D, Safavi F, Horkayne-Szakaly I, Jones R, Stram MN, Moncur JT, Hefti M, Folkerth RD, Nath A. Neurovascular injury with complement activation and inflammation in COVID-19. Brain. 2022 Jul 29;145(7):2555–2568. doi: 10.1093/brain/awac151. PMID: 35788639; PMCID: PMC9278212.

13. Solomon IH, Normandin E, Bhattacharyya S, et al. Neuropathological features of Covid-19. New England Journal of Medicine 2020; 383(10): 989992.

14. Reichard RR, Kashani KB, Boire NA, Constantopoulos E, Guo Y, Lucchinetti CF. Neuropathology of COVID-19: A spectrum of vascular and acute disseminated encephalomyelitis (ADEM)-like pathology. Acta Neuropathologica 2020; 140(1): 1–6.

15. Stram MN, Seifert AC, Cortes E, et al. Neuropathology of pediatric SARS-CoV-2 infection in the forensic setting: Novel application of ex vivo imaging in analysis of brain microvasculature. Frontiers in Neurology. 2022; 13: 894565.

16. Suh J, Mukerji SS, Collens SI, Padera RF Jr, Pinkus GS, Amato AA, Solomon IH. Skeletal muscle and peripheral nerve histopathology in COVID-19. Neurology 2021; 97(8): e849–58.

17. Maiese A, Manetti AC, La Russa R, et al. Autopsy findings in COVID-19-related deaths: a literature review. Forensic Sci Med Pathol 2021; 17(2): 279–96.

18. Stokes EK ZL, Anderson KN, et al. Coronavirus Disease 2019 Case Surveillance — United States, CDC, January 22–May 30, 2020.

19. CDC. United States COVID-19 Cases, Deaths, and Laboratory Testing (NAATs) by State, Territory, and Jurisdiction. August 3, 2022. https://covid.cdc.gov/covid-data-tracker/#cases_casesper100klast7days (accessed August 3rd, 2022).

20. World Health Organization. (2021). A clinical case definition of post COVID-19 condition by a Delphi consensus, 6 October 2021.

21. Health information of deceased individuals: Guidance portal. Health Information of Deceased Individuals | Guidance Portal. (n.d.). Retrieved September 03, 2022, from https://www.hhs.gov/guidance/document/health-information-deceased-individuals

22. Stuart, Elizabeth A. “Matching methods for causal inference: A review and a look forward.” Statistical science: a review journal of the Institute of Mathematical Statistics, vol. 25,1 (2010).

23. Imbens, G, & Rubin, D. Causal Inference for Statistics, Social, and Biomedical Sciences: An Introduction. Cambridge: Cambridge University Press (2015).

24. Bind M-AC, Rubin DB. Bridging observational studies and randomized experiments by embedding the former in the latter. Statistical Methods in Medical Research,28, 7, Pp. 1958–1978 (2019).

25. Rosenbaum P and Rubin DB. The central role of the propensity score in observational studies for causal effects, Biometrika, 70, 41–55 (1983)

26. Lee, J, Forastiere L, Miratrix L, and Pillai N. More Powerful Multiple Testing in Randomized Experiments with Non-Compliance. arXiv:1605.07242 (2016)

27. Branson Z. Randomization tests to assess covariate balance when designing and analyzing matched datasets. Observational studies 7, 1–36 (2021)

28. Wasserstein R, Schirm A, Lazar N. Moving to a World Beyond “p < 0.05”. The American Statistician, 73, 1–19 (2019)

29. DeVita MA, Wicclair M, Swanson D, Valenta C, Schold C. Research involving the newly dead: an institutional response. Crit Care Med 2003; 31(5 Suppl): S385–90.

30. Batra A, Clark JR, Kang AK, et al. Persistent viral RNA shedding of SARS-CoV-2 is associated with delirium incidence and six-month mortality in hospitalized COVID-19 patients. Geroscience 2022; 44(3): 1241–54.

31. Natarajan A, Zlitni S, Brooks EF, et al. Gastrointestinal symptoms and fecal shedding of SARS-CoV-2 RNA suggest prolonged gastrointestinal infection. Medicine (New York) 2022; 3(6): 371–87.

32. Gaebler C, Wang Z, Lorenzi JCC, et al. Evolution of antibody immunity to SARS-CoV-2. Nature 2021; 591(7851): 639–44.

33. Zollner A, Koch R, Jukic A, et al. Postacute COVID-19 is Characterized by gut viral antigen persistence in inflammatory bowel diseases. Gastroenterology 2022; 163(2): 495–506.

34. Viszlayová D, Sojka M, Dobrodenková S, et al. SARS-CoV-2 RNA in the cerebrospinal fluid of a patient with long COVID. Therapeutic Advances in Infectious Disease 2021; 8:20499361211048572.

35. Caniego-Casas T, Martínez-García L, Alonso-Riaño M, et al. RNA SARS-CoV-2 persistence in the lung of severe COVID-19 patients: A case series of autopsies. Frontiers in Microbiology 2022; 13:824967.

36. Delorey TM, Ziegler CGK, Heimberg G, et al. COVID-19 tissue atlases reveal SARS-CoV-2 pathology and cellular targets. Nature 2021; 595(7865):107–13.

